# Microscopic fractional anisotropy asymmetry in unilateral temporal lobe epilepsy

**DOI:** 10.1101/2023.05.10.23289785

**Authors:** Nico J. J. Arezza, Hana Abbas, Caroline Chadwick, Ingrid S. Johnsrude, Jorge Burneo, Ali R. Khan, Corey A. Baron

## Abstract

**Objectives:** Surgical resection is the method of choice for treating medically refractory unilateral temporal lobe epilepsy (TLE), but postsurgical prognosis depends on magnetic resonance imaging (MRI) findings. Seizure freedom is more often achieved after resection in MRI-positive patients (those with MRI abnormalities such as mesial temporal sclerosis) than in MRI-negative patients. Diffusion MRI shows promise as a marker of neuronal abnormalities due to its sensitivity to cellular changes such as axon damage, indexed by fractional anisotropy. However, fractional anisotropy is not specific to axon integrity in grey matter where axon orientation is not uniform. In contrast, microscopic fractional anisotropy is a recently introduced dMRI technique that is sensitive to axon integrity regardless of axon orientation. This work investigated whether microscopic fractional anisotropy may be sensitive to hippocampal abnormalities in unilateral TLE.

**Methods:** Diffusion MRI was performed on a 3T scanner in 9 patients (age = 33 +/- 12 years) with unilateral TLE and 9 healthy volunteers (age = 26 +/- 6). A deep learning method was employed to segment the hippocampus into smaller subfields corresponding to the subiculum, cornu ammonis (CA) 1, CA2/3, and CA4 plus dentate gyrus (DG). Mean ipsilateral and contralateral measurements of subregion volume, diffusivity, fractional anisotropy, and microscopic fractional anisotropy were compared to investigate asymmetry in each subfield.

**Results:** Microscopic fractional anisotropy was reduced, and diffusivity was elevated in the ipsilateral CA4/DG region relative to the contralateral side in all 9 patients. Asymmetries in diffusion metrics between the left and right sides of the hippocampus subfields were not observed in the healthy volunteers.

**Significance:** Diffusion MRI may complement standard imaging procedures by detecting abnormalities in MRI-negative patients. Due to its insensitivity to axon orientation, microscopic fractional anisotropy may yield a more robust measurement than fractional anisotropy and may improve epileptic focus localization in surgical candidates.

## 1. INTRODUCTION

Temporal lobe epilepsy (TLE) is the most common form of focal epilepsy in adults, with as many as two-thirds of seizure foci being localized to the temporal lobe ^1–3^. Though TLE can often be managed with anticonvulsant medications, approximately 30% of adults with epilepsy eventually develop medically intractable epilepsy despite appropriate drug therapy ^3,4^. Surgical resection of the seizure focus has been shown to be superior to medical treatment and is the method of choice for managing medically intractable TLE ^5,6^. In most of these patients, the epileptic focus lies within the mesial region of the temporal lobe and can be identified by the presence of mesial temporal sclerosis (MTS), which manifests as scarring and atrophy that can often be detected by MRI ^7^. Seizure freedom following surgical resection is achieved in 75% of patients with clearly delineated MTS in MRI (i.e. MR-positive or MR+ patients), but in only 51% of MR-negative (MR-) patients ^8^, perhaps because the seizure focus has not been adequately localized and the resection is incomplete. This demonstrates the need for highly sensitive imaging techniques to complement the current gold standard MRI, EEG, and nuclear medicine techniques, and improve seizure focus localization.

Diffusion-weighted MRI (dMRI) is a promising technique for visualizing pathological abnormalities in TLE due to its sensitivity to neuron microstructure. The diffusion tensor imaging (DTI) parameters fractional anisotropy (FA) and mean diffusivity (MD) are of particular interest because demyelination, reduced axon density, and widened extracellular spaces due to gliosis reduce FA and increase MD ^9^. Previous studies have shown that increased MD and reduced FA are present in various brain regions in TLE patients ^8–10^, and that increased MD is present in the ipsilateral side of the hippocampus in patients with unilateral TLE ^11,12^. Despite these promising results, the DTI model is inadequate for quantifying regions with crossing or fanning neuron fibers because of its sensitivity to intra-voxel fiber orientation dispersion ^13,14^. FA, in particular, significantly underestimates water diffusion anisotropy in regions with complex fiber orientations ^15^; this limits its specificity to abnormalities in TLE because the most common pathology in medically intractable TLE is hippocampal sclerosis (HS) ^16,17^ but the hippocampus contains crossing fiber regions ^18^.

Microscopic fractional anisotropy (μFA) is a recently developed dMRI metric that quantifies water diffusion anisotropy independent of both neuron fiber orientation dispersion and compartment size ^19^. Generally, μFA imaging techniques distinguish between anisotropy resulting from microstructure and anisotropy resulting from axon orientation by exploiting the contrast between two different dMRI acquisitions ^19–22^: (1) acquisitions that each probe diffusion in a single direction (i.e. encoding that is typically used in dMRI), and (2) acquisitions that probe diffusion in multiple orthogonal directions simultaneously. Previous studies have demonstrated that μFA outperforms FA for delineating lesions in multiple sclerosis ^23^, for evaluating white matter degeneration in Parkinson’s disease ^24^, and for distinguishing between different types of brain tumors ^22^, among other potential applications. In the TLE clinical workflow, μFA may provide a complementary metric to the current imaging and EEG techniques due to its sensitivity to microstructure and insensitivity to fiber orientation, particularly in brain regions containing crossing fibers, such as the hippocampus ^25^. However, the benefits of μFA imaging in TLE have not yet been assessed.

This preliminary work aims to evaluate the sensitivities of μFA, FA, MD, and regional volume to detect abnormalities in four hippocampal subregions in patients with unilateral TLE. Asymmetries in measurements of anisotropy, diffusivity, and volume between the ipsilateral and contralateral hemispheres may indicate unilateral abnormalities that can lateralize the epileptic focus. We hypothesize that μFA may be more sensitive to hippocampal abnormalities than FA due to its independence from neuron fiber orientation and may usefully complement the current standard-of-care for diagnostic or pre-surgical imaging in TLE.

## 2. METHODS

### 2.1 Participants

Nine TLE patients (four female and five male, mean age ± standard deviation = 33 ± 12 years) and nine healthy volunteers (four female and five male, mean age ± standard deviation = 26 ± 6 years) were recruited for this study, which was approved by the health sciences research ethics board at Western University. Informed consent was obtained from all participants prior to their recruitment. The following inclusion criteria were used to determine eligibility for the TLE cohort: all patients (a) had a history of epilepsy, (b) underwent radiological and/or comprehensive EEG assessments to identify and lateralize the epileptogenic region, and (c) were suspected to have a unilateral seizure focus in the temporal lobe. Three patients in the TLE cohort underwent unilateral temporal lobectomy after imaging and post-surgical pathology confirmed the presence of MTS; they are herein referred to as the “confirmed MTS” subgroup, while the other six patients are referred to as the “MR-negative” subgroup. Clinical and demographic information for the patient participants is shown in Table 1.

**Table 1.** Clinical characteristics of patients with left and right temporal lobe epilepsy.

| Age | Sex | Handedness | Scalp EEG | Intracranial EEG | Ipsilateral Side | 1.5T MRI Radiological Findings (T1 and T2) | Post-Surgical Pathology |
| --- | --- | --- | --- | --- | --- | --- | --- |
| 40-44 | M | R | L TLE | N/A | L | MTS | MTS+ |
| 30-34 | F | R | L TLE | N/A | L | MTS | MTS+ |
| 20-24 | M | R | L TLE | N/A | L | MTS | MTS+ |
| 55-59 | M | R | L TLE | N/A | L | Smaller left hippocampus compared to right | N/A |
| 20-24 | F | R | L TLE | L TLE | L | N/A | N/A |
| 30-34 | F | R | L TLE | L TLE | L | 1.2 cm possible polyp in nasal cavity | N/A |
| 35-39 | F | R | R TLE | R TLE | R | Chronic mucosal thickening in the paranasal sinuses | N/A |
| 25-29 | M | L | R TLE | N/A | R | Apparent cyst near right lateral ventricle with stable appearance | N/A |
| 25-29 | M | R | L TLE | N/A | L | Slightly thicker cortex and less myelination in left temporal pole compared to right | N/A |

**EEG**-Electroencephalogram, **L**-Left, **R**-Right, **TLE**-Temporal Lobe Epilepsy, **N/A**-Not Available, **MTS**-MTS Detected, **MTS+**-MTS Confirmed Surgically

### 2.2. MRI acquisition and processing

Participants were scanned using a 3T full-body MRI system (Siemens Prisma) with a 32-channel head coil. The protocol consisted of two anatomical MRI scans followed by two dMRI scans for separate DTI and μFA acquisitions. The first anatomical scan was a T1-weighted magnetization-prepared rapid acquisition with gradient echo (MPRAGE) sequence with repetition time/echo time (TE/TR) = 2.3/2400ms and inversion time = 1.06s, and the second anatomical scan was a T2-weighted sequence with TE/TR = 564/3200ms. Both the T1- and T2-weighted scans had a field-of-view (FOV) = 240x256mm^2^, 0.8mm isotropic voxel size, and used rate 2 generalized auto-calibrating partially parallel acquisitions (GRAPPA). The DTI scan used a multiband echo-planar imaging (EPI) sequence with TE/TR = 99/5500ms, rate 2 GRAPPA, FOV = 222x222mm^2^, and 1.6mm isotropic voxel size to acquire 6, 36, and 60 linear tensor-encoded (LTE) volumes at b-values of 0, 1000, and 2000s/mm^2^, respectively, with a total scan time of 9 minutes. The μFA scan used a multiband EPI sequence with TE/TR = 92/4900ms, rate 2 GRAPPA, FOV = 229x229mm^2^, and 1.8mm isotropic voxel size to acquire 8 LTE volumes at b=2000s/mm^2^ and 3, 6, and 16 spherical tensor-encoded (STE) volumes at b=100, 1000, and 2000s/mm^2^, respectively, with a total scan time of 3 minutes. The μFA scan was performed twice, first with anterior-to-posterior and then with posterior-to-anterior phase encoding directions. Principal component analysis denoising and Gibbs’ ringing artifact correction were performed on the dMRI volumes with the DWIDENOISE ^26,27^ and MRDEGIBBS ^28^ toolboxes from Mrtrix3 ^29^ and the data were then corrected for EPI readout and eddy current distortions using TOPUP ^30^ and EDDY ^31^ from FSL ^32^.

### 2.3 Hippocampus segmentation

A deep-learning surface-based hippocampus unfolding pipeline (*Hippunfold v0*.*5*.*1* ^33^) was used to segment the hippocampus into subiculum (SB), cornu ammonis (CA) 1-4, and dentate gyrus (DG) subfields, using the T2-weighted volume as input. The volumetric subfield segmentations from HippUnfold were used in this study, which are generated by 1) segmentation of hippocampal tissue and external boundaries with a U-net model, 2) mapping the intrinsic coordinates of the hippocampal gray matter using Laplace’s equation, and 3) transferring subfield boundaries to each individual tissue segmentation using the unfolded coordinates and a subfield atlas defined from a 3D histology reference space (labelling CA1,CA2,CA3,CA4, subiculum, and dentate gyrus). To reduce the number of comparisons during analysis, some of the subfields were combined to form distinct subregions based on the following three International League Against Epilepsy (ILAE) histopathological HS classifications: HS ILAE Type 1 is defined as severe neuron loss and gliosis primarily in CA1 and CA4; in Type 2 loss and gliosis predominate in CA1; and in Type 3 they predominate in CA4 ^17^. Although significant cell loss is observed in CA2 and/or CA3 in some TLE patients, these findings are not consistent across any of the HS ILAE types ^17^ so these adjacent regions were combined into one subregion. The CA4 subfield was combined with the adjacent DG since cell loss scores in the DG tend to be higher in CA4-predominant HS type 1 and type 3 than in type 2 ^17^. The CA1 subfield was not merged with any others as it is of interest in HS type 1 and type 3. The hippocampus subregions and their relation to the ILAE histopathological HS types are summarized in Table 2.

**Table 2.** Hippocampus subregions and their significance in hippocampal sclerosis histopathology.

| *Subregion* | *Significance in hippocampal sclerosis histopathology* |
| --- | --- |
| **SB** | Cells are often (but not always) preserved in the subiculum. |
| **CA1** | Severe neuron loss and gliosis occurring in CA1 is a hallmark of ILAE HS types 1 and 2. |
| **CA2/3** | Cell loss scores in CA2 and/or CA3 are highly variable across patients with HS. |
| **CA4/DG** | Severe neuron loss and gliosis occurring in CA4 is a hallmark of ILAE HS types 1 and 3.<br>Cell loss scores in the DG also tend to be higher in ILAE HS types 1 and 3 than in type 2. |

### 2.4 Estimation of dMRI parameters

To ensure that all dMRI metrics were mapped to the same coordinate system, the DTI volumes were registered to the μFA image space using the linear registration tool FLIRT ^34^ from FSL. MD and FA maps were computed by fitting the dMRI data with b≤1000s/mm^2^ from the DTI scan to the DTI model using a weighted linear least-squares method ^35,36^. μFA maps were computed by performing a joint fit between the entire set of LTE and STE data from the μFA scan to the second order cumulant model as described by Arezza *et al* ^37^. The T1-weighted image volumes were registered to the μFA space and then the inverse transformations were used to register the MD, FA, and μFA maps to the anatomical space. To ensure good registration quality, outlines of the hippocampal subregions were overlaid on top of the registered MD, FA, and μFA maps and were visually inspected.

### 2.5 Statistical analysis

For each TLE patient, the mean MD, FA, and μFA were measured in the ipsilateral and contralateral sides of each of the four hippocampal subregions and full hippocampus, and the volume of each subregion was measured by computing the sum of the number of voxels in the region. The mean and standard deviation of each of the four measurements of interest, across all patients, were computed for the ipsilateral and contralateral sides. In the healthy volunteer cohort, the same measurements were made in the left and right sides for each subfield, as well as average measurements spanning both sides of the brain. For each metric in each subregion, a paired t-test was performed to test for significant differences between the ipsilateral and contralateral sides in the TLE cohort, and an unpaired t-test was performed to test for significant differences between the ipsilateral side of the TLE group and the average of the left and right sides in the healthy group. The Bonferroni correction was applied to the p-values to account for multiple comparisons.

To quantify asymmetries between the two hemispheres, the percentage differences between the ipsilateral and contralateral measurements in the TLE group were computed in each subregion and in the whole hippocampus for each patient using the following equation:

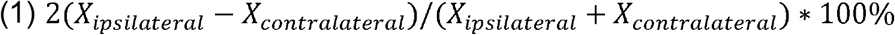

where *X* is the measurement of interest. It was hypothesized that volume, FA, and μFA may be reduced, and MD may be elevated, in some ipsilateral regions compared to their respective contralateral counterparts due to tissue atrophy, gliosis, and changes to microstructure. Notably, asymmetries may be more likely to be observed in the CA1 and CA4/DG subregions that are predominantly affected in HS than in the SB and highly variable CA2/3 subregions. For the healthy group, asymmetry was measured within subregions and in the whole hippocampus by comparing the left and right sides.

## 3. RESULTS

Example sagittal and coronal T1- and T2-weighted images from one of the healthy volunteers are depicted in Fig. 1 with the four hippocampal subregions outlined. All hippocampal segmentations were manually inspected for accuracy in delineating the hippocampal tissue and subfields. Coronal slices of T1-weighted MRI, MD, FA, and μFA from a TLE patient with confirmed MTS are depicted in Fig. 2 for comparison. Ipsilateral and contralateral measurements of volume (normalized against the mean contralateral volume), MD, FA, and μFA are plotted in Fig. 3 for all subregions, in addition to average measurements spanning both the right and left side for all subregions in healthy volunteers. Notably, MD was significantly elevated and μFA was significantly reduced in the ipsilateral CA4/DG region relative to the contralateral side in TLE patients, with Bonferroni-corrected p-values of 0.048 and 0.018, respectively. Compared to the average values in the healthy cohort, ipsilateral MD was significantly elevated in every subregion except CA1, and ipsilateral μFA was significantly reduced in every subregion. Although the mean ipsilateral volume was reduced relative to the contralateral side in all four subregions and in the full hippocampus in the TLE cohort, this metric varied considerably from patient to patient and the difference was not statistically significant in any region. However, ipsilateral volume was significantly reduced in every region except SB relative to average measurements in the healthy cohort. For the FA metric, no significant asymmetries were observed in the patients in any of the subregions.

**Figure 1.**
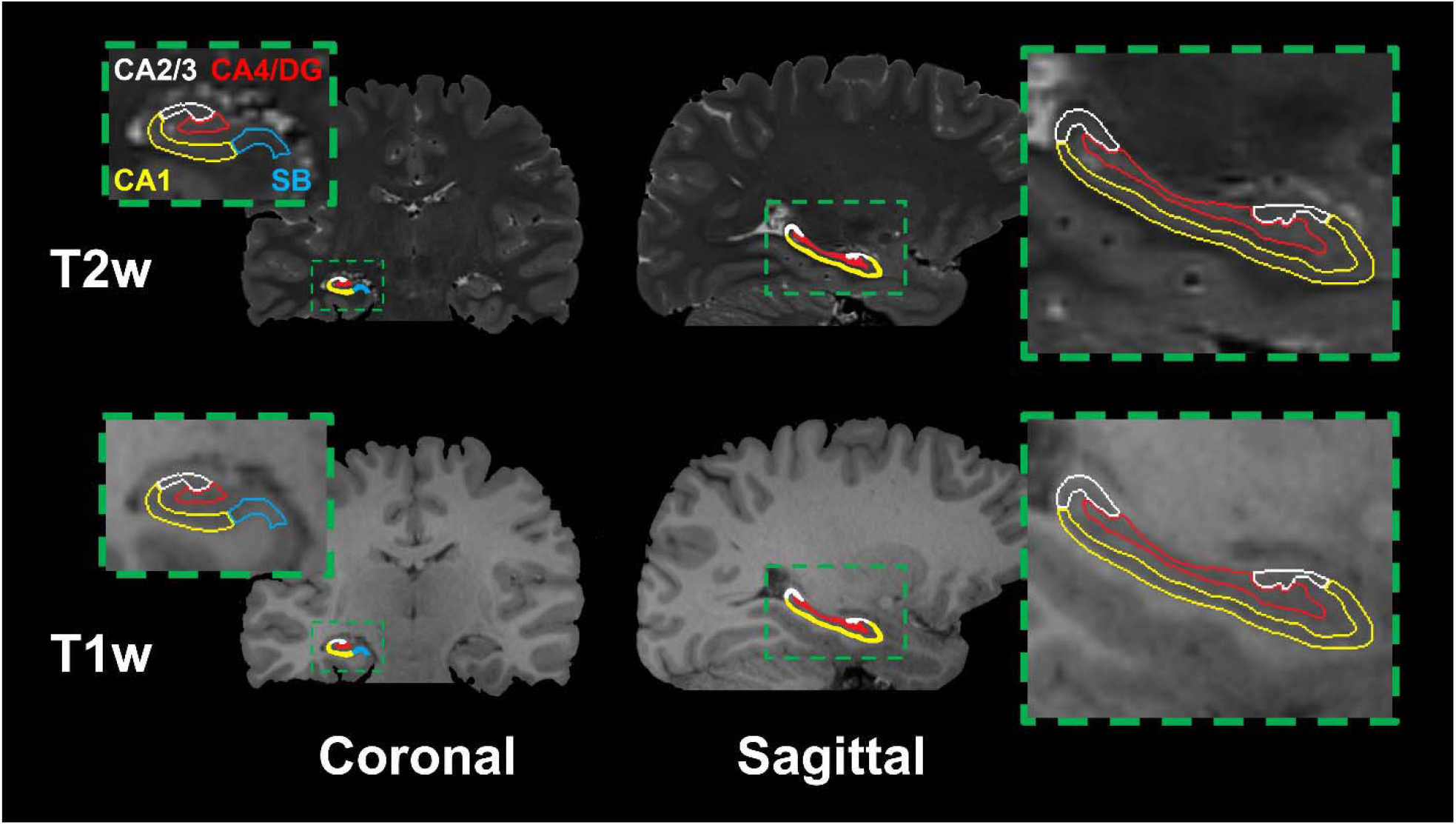
Sagittal and coronal T1-weighted (top) and T2-weighted (bottom) MR images from a healthy volunteer with insets highlighting the four hippocampal subregions used in this study: the subiculum (SB), cornu ammonis 1 (CA1), cornu ammonis 2 and 3 (CA2/3) and cornu ammonis 4 plus dentate gyrus (CA4/DG). Note that only the right hippocampus is labeled although both hippocampi were analyzed.

**Figure 2.**
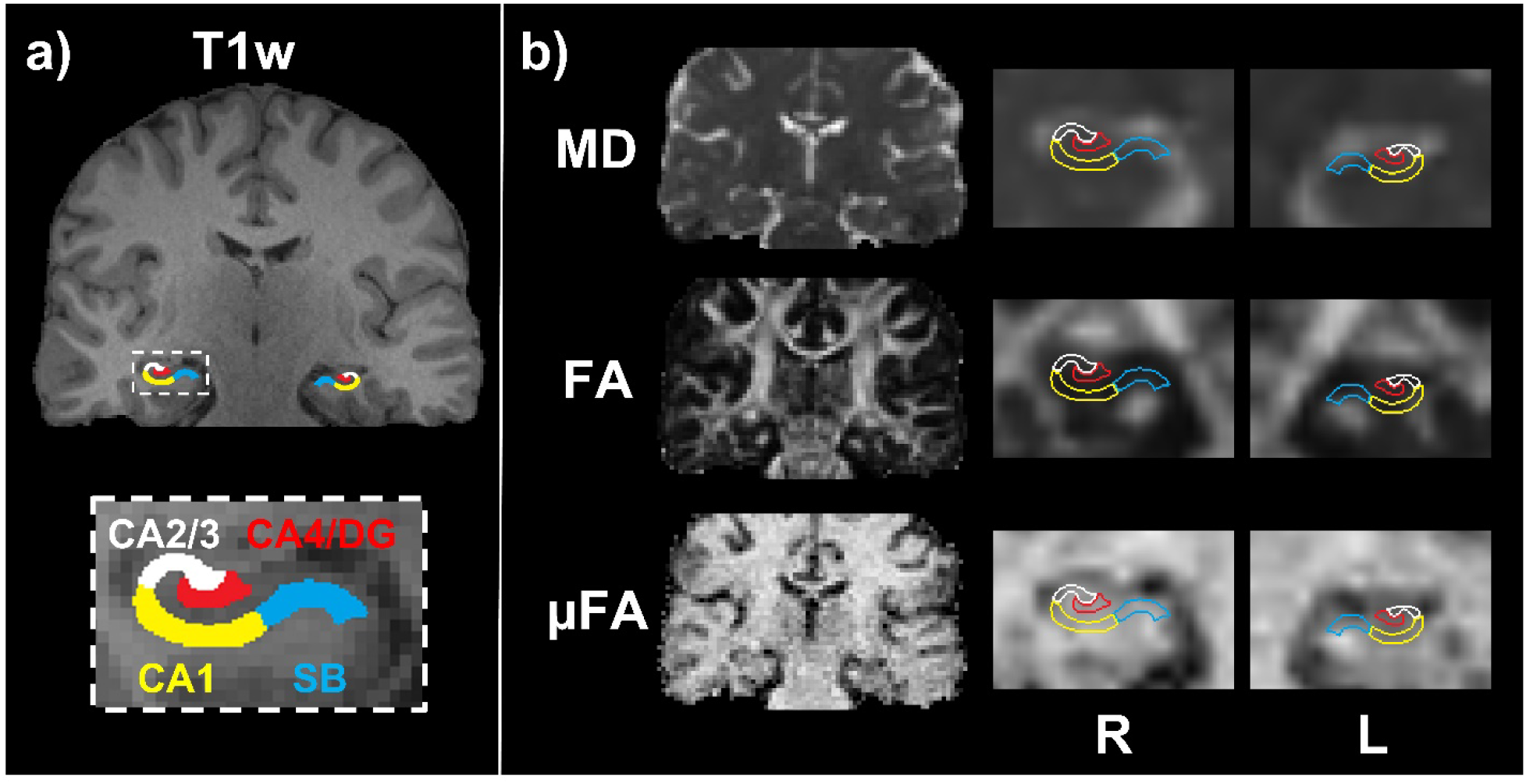
(a) Example T1-weighted coronal image from a TLE patient with confirmed MTS, with the four hippocampal subregions highlighted. (b) MD, FA, and μFA coronal slices from the same patient before registration to T1-space (left), and after registration to T1-space and interpolation (right) depicting the ipsilateral and contralateral hippocampal regions. Note that for this patient, the left side (L) is the ipsilateral side and the right side (R) Is the contralateral side.

**Figure 3.**
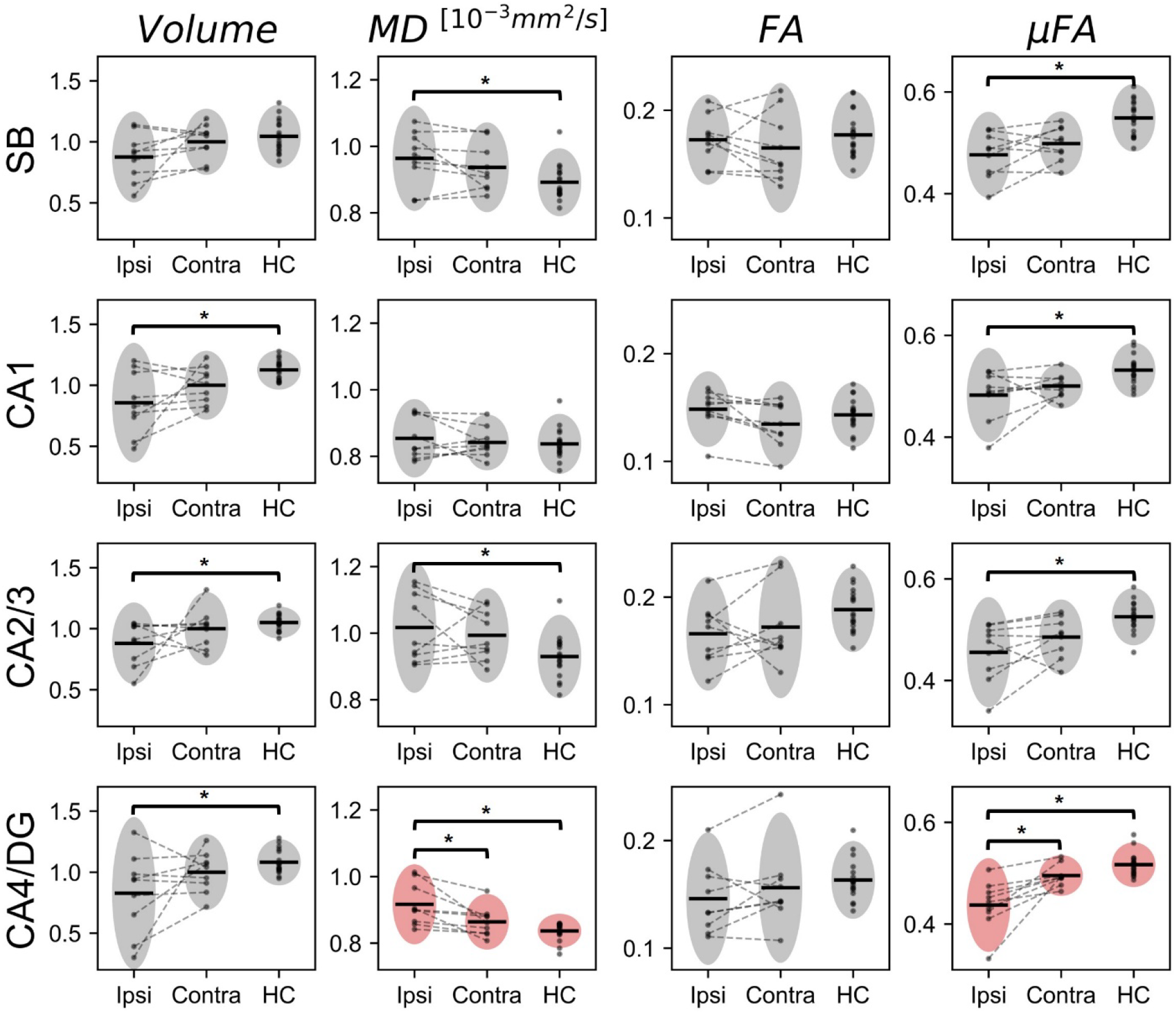
Volume (normalized against the mean contralateral volume), MD, FA, and μFA measurements in the ipsilateral and contralateral sides of each of the four hippocampal subregions across 9 TLE patients, plus mean measurements of both hemispheres across 9 healthy control volunteers (HC). The horizontal black lines depict the mean measurement across the cohort, and the gray ovals highlight a region spanning two standard deviations above and below the mean. The two plots with pink ovals highlight significant MD (p=0.048) and μFA (p=0.018) asymmetries in the TLE cohort.

To further investigate asymmetries in the CA4/DG subregion, and to compare these asymmetries with full-hippocampus measurements, the percentage differences between ipsilateral and contralateral measurements in CA4/DG and in the full hippocampus were plotted in Fig. 4. Overall, the mean percentage difference between the ipsilateral and contralateral measurements in the CA4/DG region was -24.4% for volume, +5.8% for MD, -6.6% for FA, and -12.9% for μFA; in the full hippocampus the mean percentage difference between ipsilateral and contralateral measurements was - 16.5% for volume, +2.7% for MD, +3.3% for FA, and -5.9% for μFA.

**Figure 4.**
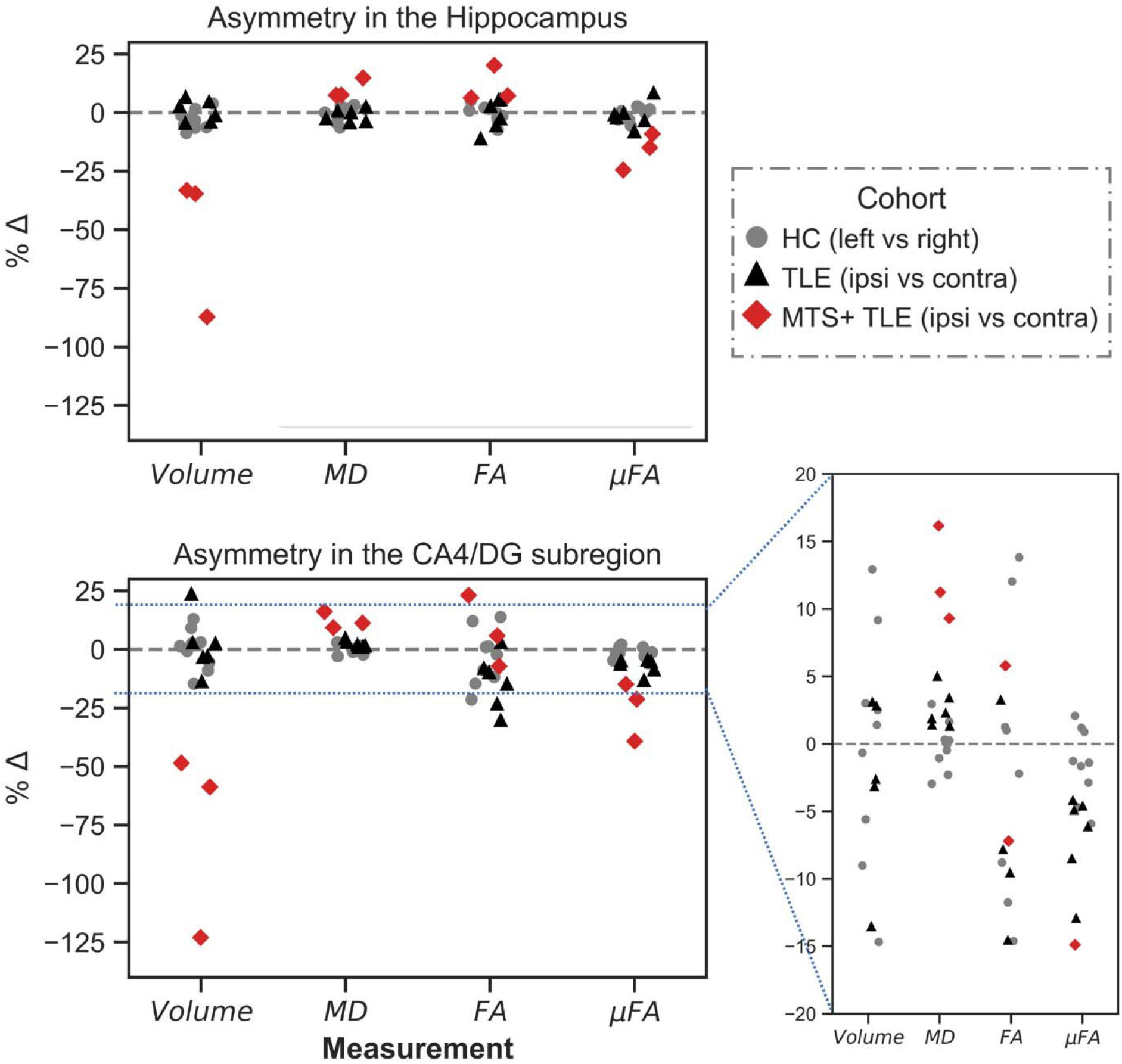
Percentage difference (%Δ) between ipsilateral and contralateral measurements of volume, MD, FA, and μFA in the full hippocampus (top) and CA4/DG region (bottom) in unilateral TLE patients, and %Δ between left and right measurements in 9 healthy control volunteers (HC).

All three patients with confirmed MTS were found to have reduced volume and μFA and increased MD in both the full hippocampus and the ipsilateral CA4/DG subregion relative to the contralateral side. Only one of these patients had reduced ipsilateral FA in CA4/DG, and none had reduced ipsilateral FA in the full hippocampus. Of the six MR-negative patients, only three had reduced volume in ipsilateral CA4/DG and in the full hippocampus relative to the contralateral side. Five had reduced ipsilateral FA in CA4/DG and four had reduced ipsilateral FA in the full hippocampus. All six MR-patients had reduced μFA and increased MD in the ipsilateral CA4/DG subregion relative to the contralateral side, but only five had reduced μFA and increased MD in the ipsilateral full hippocampus. Generally, greater asymmetries were observed between hemispheres in the CA4/DG subregion than across the entire hippocampus in the TLE cohort. Overall, the mean percentage difference between the ipsilateral and contralateral measurements in the CA4/DG region was -24.4% for volume, +5.8% for MD, -6.6% for FA, and -12.9% for μFA; in the full hippocampus the mean percentage difference between ipsilateral and contralateral measurements was -16.5% for volume, +2.7% for MD, +3.3% for FA, and -5.9% for μFA.

## 4. DISCUSSION

In this preliminary study, the dMRI metrics of MD, FA, and μFA were measured in several hippocampal subfields, and compared to a volumetric measurement, to assess whether they demonstrate sensitivity to unilateral hippocampal abnormalities in TLE patients. This study is the first to apply μFA imaging to the study of TLE. It was observed that MD was significantly elevated and μFA was significantly reduced in the ipsilateral CA4/DG region, relative to the contralateral side, in all patients. This subregion is affected by severe cell loss and gliosis in TLE patients with ILAE HS types 1 and 3. The increased ipsilateral MD is consistent with other diffusion MRI studies of temporal lobe epilepsy ^11,38–41^. In particular, Goubran *et al* observed a strong negative correlation between MD and cell density in CA4/DG ^12^. μFA values were more asymmetric (between hemispheres) than were MD values in the CA4/DG region, suggesting that it is more sensitive to hippocampal abnormalities. Although the mean CA4/DG volume asymmetry across patients was greater than those of the dMRI metrics, a decreased ipsilateral volume in the region correctly predicted the side of the epileptic focus in only six of nine patients, while MD and μFA measurements demonstrated asymmetry in the CA4/DG that was consistent with the EEG results in all nine patients.

We hypothesize that the reduced ipsilateral μFA stemmed from the loss of axons that invariably occurs when neurons die. Notably, axons are more sensitive to homeostatic imbalances than cell bodies, and so are generally lost earlier: when under stress axons can degenerate while the cell body remains ^42–45^. Accordingly, these results, although from a small sample, suggest that μFA may be an early marker of mesial temporal sclerosis.

### 4.1 Confirmed MTS vs. MR-negative TLE

Both full hippocampus and CA4/DG-specific asymmetries in region volume, MD, and μFA correctly lateralized the epileptic focus in all three patients with confirmed MTS. These patients exhibited considerable unilateral hippocampal atrophy which, when combined with concordant results from EEG and other clinical testing, made them candidates for anterior temporal lobectomy procedures. All three patients with confirmed MTS had total hippocampal volume asymmetries greater than 30% (prior to surgery) and CA4/DG volume asymmetries greater than 45%, while the six MR-negative patients had total hippocampal volume asymmetries of <10% and CA4/DG volume asymmetries of <20%. The confirmed MTS patients also had the greatest MD and μFA asymmetries.

The inability of volume asymmetries to lateralize the epileptogenic zone in the MR-negative cohort highlights the need for supplementary imaging techniques in the TLE clinical workflow. In the MR-subgroup, the full hippocampus and CA4/DG volume asymmetries correctly lateralized the epileptic focus in only half of the patients. Right-left hemispheric asymmetry of hippocampal volume occurs in healthy subjects and is not necessarily indicative of pathology or injury ^46^. In contrast, diffusion metrics are linked to microstructural changes that suggest neuron damage or gliosis, perhaps giving said metrics better specificity to unilateral hippocampal abnormalities relevant to TLE. The results of this work support this theory as MD and μFA asymmetries in CA4/DG correctly lateralized the epileptic focus for all six MR-negative patients, regardless of whether the ipsilateral side was in the left or right hemisphere, and regardless of whether CA4/DG volume was reduced or elevated in that side.

### 4.2 Microscopic fractional anisotropy vs. fractional anisotropy

Since both FA and μFA index water diffusion anisotropy, they are particularly comparable. Mean FA values typically fell in the 0.1-0.2 range across all hippocampal subregions and were consistently lower than mean μFA values, which fell in the 0.4-0.5 range. This discrepancy likely resulted from crossing and fanning fibers in the hippocampus, which attenuate FA measurements but do not affect μFA. The mean values for both anisotropy metrics were consistent with the results of Yoo *et al* ^25^, in which mean FA and μFA values of 0.2 and 0.47, respectively, were observed in the hippocampi of healthy volunteers.

Although CA4/DG was the only region in which a statistically significant asymmetry in μFA was observed in the TLE cohort, mean ipsilateral μFA was consistently reduced relative to both the contralateral and average healthy control μFA across all four hippocampal subregions (Fig. 3). FA asymmetry in the TLE group was not statistically significant in any of the subregions and was inconsistent across regions. Given that FA values were significantly lower in all subregions relative to μFA values, and that no significant FA asymmetries were observed, it is likely that the sensitivity of FA in detecting hippocampal abnormalities in TLE is suppressed by its lack of specificity to neuron fiber microstructure and that μFA is a more suitable measure of diffusion anisotropy in brain regions containing crossing fibers.

### 4.3 Limitations

This preliminary work was limited by the small size of the unilateral TLE patient cohort. Although μFA and MD measurements were reliably asymmetric in CA4/DG, future work should include larger patient cohorts to validate these findings and to potentially elucidate asymmetries in other subregions, such as CA1.

Since ILAE HS type 1 is the most common subtype of HS, accounting for 60-80% of TLE-HS cases ^17,47,48^, it was expected that asymmetries might present in the CA1 and CA4/DG regions. However, no significant asymmetries were observed in CA1 in any of the metrics. It may be the case that MD and μFA are more sensitive to abnormalities in the CA4/DG region, but the small sample size may have affected the results.

The spatial resolutions of the dMRI volumes acquired in this study (1.8 mm isotropic for μFA) are suboptimal for visualizing hippocampal subfields ^49^, so some partial volume effects near the boundaries between subregions and near CSF likely affected the results. Since the SB, CA1, and CA2/3 subregions encompass the periphery of the hippocampus, they could be more susceptible to partial volumes of extra-hippocampal brain tissue or CSF; contrarily, the CA4/DG region lies in the center of the hippocampus and would only be affected by partial volumes of other hippocampal subregions. The significant asymmetries in MD and especially μFA in CA4/DG demonstrate the potential for dMRI in lateralizing the epileptic zone in TLE, and demonstrate the increased utility of μFA over FA in studying the hippocampus. In future work, the spatial resolution could be improved at the expense of increased scan duration; the μFA protocol used in this study required 3 minutes to achieve 1.8 mm resolution, but Yoo *et al* demonstrated a μFA protocol with 1.5 mm isotropic resolution that could be acquired in 15 minutes ^25^. To counter the increased scan time needed for higher resolution, the field-of-view and number of slices could be reduced to capture a smaller subvolume of the brain containing the hippocampus. Additionally, techniques to mitigate CSF partial-volume effects, such as a recently proposed free water elimination μFA protocol ^50^, could be employed.

## 5. Conclusions

This study demonstrated that the combination of hippocampal subfield segmentation with μFA and MD imaging may be helpful for lateralizing the epileptogenic zone in patients with unilateral TLE. Assuming the poorer surgical outcomes experienced by patients with MR-TLE are in part due to poorer identification of the epileptic focus, then dMRI techniques that can complement the current techniques for lateralizing and localizing the epileptic focus may be able to improve surgical outcomes in these patients.

Both the DTI and μFA protocols in this work are clinically feasible and could easily be included in a clinical workflow, as both scans were performed at a clinical field strength of 3T and each only required 6 minutes or fewer of total scan time (as the b=2000s/mm^2^ acquisitions in the DTI scan were redundant). To further optimize the protocol, MD and FA could be estimated from a μFA scan by fitting the low b-value data (<1000s/mm^2^) to the diffusion tensor model, eliminating the need for a separate DTI scan, though this was not possible in this work because only STE scans were acquired at the lower b-values.

## Data Availability

Data are not available due to reasons of sensitivity.

## Acknowledgements

This work was supported by the Canada Research Chairs Program (**CB, AK**), Canada First Research Excellence Fund to BrainsCAN, a Natural Sciences and Engineering Research Council (NSERC) Discovery grant (**IJ**), a Canadian Institutes of Health Research (CIHR) project grant (**AK**), a CIHR operating grant (**IJ**), and the NSERC Canada Graduate Scholarship Doctoral (CGS-D) program (**NA**).

